# Early Hyperoxaemia and 2-year Outcomes in Infants with Hypoxic-ischemic Encephalopathy- a Secondary Analysis of the Infant Cooling Evaluation (ICE) trial

**DOI:** 10.1101/2023.08.25.23294627

**Authors:** Shiraz Badurdeen, Jeanie L Y Cheong, Susan Donath, Hamish Graham, Stuart B Hooper, Graeme R Polglase, Sue Jacobs, Peter G Davis

## Abstract

**Objective(s):** To determine the causal relationship between exposure to early hyperoxaemia and death/disability in infants with hypoxic-ischemic encephalopathy (HIE).

**Study design:** We analyzed data from the Infant Cooling Evaluation (ICE) trial that enrolled newborns ≥35 weeks’ gestation with moderate-severe HIE, randomly allocated to hypothermia or normothermia. The primary outcome was death or major sensorineural disability at 2 years. We included infants with arterial pO_2_ measured within 2 h of birth. Using a directed acyclic graph, we established that markers of severity of perinatal hypoxia-ischemia and pCO_2_ were a minimally sufficient set of variables for adjustment in a regression model to estimate the causal relationship between arterial pO_2_ and death/disability.

**Results:** Among 221 infants, 116 (56%) had arterial pO_2_ and primary outcome data. The unadjusted analysis revealed a U-shaped relationship between arterial pO_2_ and death/disability. Among hyperoxaemic infants (pO_2_ 100–500 mmHg) the risk of death/disability was 40/58 (0.69), while the risk in normoxaemic infants (pO_2_ 40 – 99mmHg) was 20/48 (0.42). In the adjusted model, hyperoxaemia increased the risk of death/disability (adjusted risk ratio 1.61, 95% CI 1.07 – 2.00, p= 0.03) in relation to normoxaemia.

**Conclusions:** Early hyperoxaemia increased the risk of death/disability among infants who had an early arterial pO_2_ in the ICE trial. Limitations include the possibility of residual confounding and other causal biases. Further work is warranted to confirm this relationship in the era of routine therapeutic hypothermia.

## Introduction

The past two decades have brought into focus the potential dangers of hyperoxia in critically ill patients across age spectrum.^1,2^ Studies in term newborns were among the first to provide clinical evidence in this regard; quasi-randomized trials found lower mortality when resuscitation commenced in room air versus 100% oxygen.^3^ These studies evaluated newborns with mild degrees of perinatal hypoxic-ischemia who predominantly received respiratory support alone in the delivery room. At the more severe end of the spectrum of perinatal hypoxic-ischemia there are scarce data on the effect of oxygen exposure for term infants.^4^ Despite the introduction of therapeutic hypothermia in high-resource settings, these infants continue to suffer a 40-50% risk of death or moderate-to-severe disability.^5^ Across the world, birth asphyxia is the second largest contributor to neonatal mortality.^6^ As availability of oxygen grows in lower resourced settings,^7^ the question of whether hyperoxia may cause harm in infants with perinatal hypoxic-ischemia has become increasingly important.

Recommendations regarding supplemental oxygen use for infants with perinatal hypoxic-ischemia are limited. Following the commencement of resuscitation in room air, guidelines suggest increasing the fraction of inspired oxygen to 100% if chest compressions are commenced.^8^ Once the circulation is restored, fraction of inspired oxygen (FiO_2_) is typically titrated to target oxygen saturations levels (SpO_2_) that have been observed in well infants during healthy fetal-to-neonatal transition. This strategy may result in excess oxygen delivery relative to cerebral oxygen consumption.^9^ Following admission to neonatal intensive care, it is typical to continue to target SpO_2_ of 95-98%. Recent data suggests that this oxygen saturation target results in a 30% likelihood of hyperoxaemia (pO_2_ >99 mmHg) among mechanically ventilated term infants.^10^

Whether exposure to hyperoxaemia in the first few hours after resuscitation contributes to multi-organ injury is unknown. This question has recently garnered significant interest in pediatric and adult intensive care settings.^11,12^ To explore this question in relation to term infants with hypoxic-ischemic encephalopathy (HIE) and generate preliminary clinical data, we used the dataset from the Infant Cooling Evaluation (ICE) randomized trial of therapeutic hypothermia.^13^ We aimed to evaluate whether exposure to hyperoxaemia following resuscitation was causally related to death or moderate-to-severe disability compared to normoxaemia.

## Methods

The Research Ethics Office of the Royal Children’s Hospital, Melbourne, approved the secondary use of participant data from the ICE trial for this analysis (Reference: QA/97681/RCHM-2023). The ICE trial was a randomized controlled trial conducted at 28 neonatal intensive care units in Australia, New Zealand, Canada, and the United States between 2001 and 2007.^13^ The trial was approved by the human research and ethics committee at each participating site. Participants provided written informed consent. Newborns of 35 weeks’ gestation or more were eligible if they had evidence of moderate or severe encephalopathy based on modified Sarnat criteria and indicators of peripartum hypoxia-ischemia. Infants were excluded if hypothermia could not start within 6 hours of birth, if the birth weight was less than 2 kg, if major congenital abnormalities were suspected, if there was overt bleeding, if the infant required more than 80% inspired oxygen, if death was imminent (refractory hypotension or acidosis unresponsive to treatment), or if therapeutic hypothermia had commenced before assessment.

Infants were randomized to either whole-body hypothermia to 33.5°C for 72 hours or normothermia. The primary composite outcome was death or major sensorineural disability at 2 years of age. Major sensorineural disability comprised neuromotor delay (cerebral palsy [CP] in which the child was not walking [moderate CP] or was unlikely to walk [severe CP] at 2 years, a Psychomotor Development Index score on the Bayley Scales of Infant Development II [BSID-II] of less than −2 SDs, a Motor Composite Scale score on the BSID-III of less than −2 SDs, or a disability level on the Gross Motor Function Classification System [GMFCS] of 2-5), developmental delay (a Mental Development Index score on the BSID-II of less than −2 SDs or a Cognitive Scale score or a Language Composite Scale score on the BSID-III of less than −2 SDs), blindness, and/or deafness requiring amplification or worse.

### Participants

We included all infants in the ICE trial who had an arterial blood pO_2_ measured within 2 hours of birth. This time cut-off was selected as the exposure of interest was hyperoxaemic exposure following resuscitation and most arterial blood samples were taken within this timeframe.

### Analysis

We plotted a locally estimated scatterplot smoothing (loess) curve to describe the unadjusted, univariable relationship between arterial pO_2_ and the probability of death or disability (*R* V.3.6.2, *R* Foundation, Vienna, Austria). For the adjusted analysis, we considered variables that may be associated with both the exposure and the outcome based on clinical experience and previous studies.^14–16^ To characterize the relationships between variables, we created a directed acyclic graph (DAG) using the Dagitty web-based application.^17^ We used the DAG to establish a minimally sufficient adjustment set of variables needed to estimate the causal relationship between early hyperoxaemia and death or disability.

Given the non-linear univariable relationship between arterial pO_2_ and the study outcome, we dichotomized the exposure to normoxaemia (40-99mmHg) and hyperoxaemia (100-500mmHg).^18^ Infants with pO_2_ <40 mmHg (hypoxaemic range, n= 5) and extreme outliers for pO_2_ (>500 mmHg, n= 5) were excluded from the model. Based on the adjustment variables identified in the DAG, we performed log-binomial regression to calculate the adjusted estimate of the causal effect of hyperoxaemia on the outcome of death/disability. We report the adjusted risk ratio, 95% confidence intervals and p-value, with a pre-specified statistical significance threshold of p< 0.05. We considered exploring whether the effect estimate of hyperoxaemia on death/disability was modified by the randomly allocated trial intervention (cooling). However, interaction/subgroup analyses were considered inappropriate given the lack of statistical power with the relatively small number of participants for this analysis.

## Results

Among the 221 infants in the ICE trial, n= 121 (55%) had an arterial blood gas pO_2_ available within 2h of birth. Five of these infants (4%) had no data for the primary outcome. As this proportion was small, these infants were excluded from the analysis. The baseline characteristics of included infants (n=116) are shown in table 1. Seventy-eight percent of infants had moderate or severe encephalopathy and 52% received whole-body hypothermia. Death or major sensorineural disability at 2 years of age occurred in 66 (57%). Thirty-five (30%) infants died.

**Table 1.** Baseline characteristics of infants. *Among 22 infants with an arterial blood gas who were excluded, 4 had no pO_2_ value, 13 were sampled at >2 h after birth, and 5 had no data for the primary outcome.

| Characteristic | All infants with an arterial $pO_2$ within 2h of birth ($n = 116$) | Excluded infants ($n=105$) |
| --- | --- | --- |
| *Intrapartum complications* |  |  |
| Cord prolapse | 10 (9%) | 11 (10%) |
| Shoulder dystocia | 10 (9%) | 13 (12%) |
| Antepartum haemorrhage | 23 (20%) | 17 (16%) |
| Other sentinel event | 31 (27%) | 32 (30%) |
| Caesarean birth | 59 (51%) | 47 (45%) |
| *Neonatal* |  |  |
| Gestation, weeks, mean (SD) | 39.1 (1.7) | 39.1 (1.8) |
| Birth weight, g, mean (SD) | 3348 (588) | 3523 (621) |
| Male | 63 (54%) | 64 (61%) |
| Birth outside a tertiary maternity hospital | 63 (54%) | 71 (68%) |
| Apgar score, median (IQR) |  |  |
| 1 min | 1 [0 – 2] | 1 [0 – 2] |
| 5 min | 3 [1 – 4] | 3 [1 – 4] |
| 10 min | 4 [3 – 5], ($n = 115$) | 4 [3 – 5], ($n = 99$) |
| *Resuscitation* |  |  |
| Ventilation | 116 (100%) | 105 (100%) |
| Chest compressions | 73 (63%) | 65 (62%) |
| Epinephrine | 52 (45%) | 41 (39%) |
| *Peripartum hypoxia-ischemia* |  |  |
| Apgar score at $\leq 5$ at 10 min | 101 (87%), ($n = 115$) | 80 (81%), ($n = 99$) |
| Resuscitation or ventilation at $\geq 10$ min | 114 (100%) | 98 (93%) |
| Cord or gas within 2 h of birth, pH, mean (SD) | 6.92 (0.22), (n=114) | 6.92 (0.22), (n= 97) |
| *Blood gas route* |  |  |
| Arterial | 116 (100%) | 22 (22%)* |
| Capillary |  | 38 (37%) |
| Venous |  | 42 (41%) |
| Unknown |  | 3 (3%) |
| *Arterial pO<sub>2</sub> (mmHg)* |  |  |
| <40 | 5 (4%) | 1 (5%) |
| 40 – 99 | 48 (41%) | 8 (36%) |
| ≥100 | 63 (54%) | 9 (41%) |
| Unknown | 0 (0%) | 4 (18%) |
| *Encephalopathy at assessment* |  |  |
| Mild | 23 (20%) | 19 (18%) |
| Moderate | 61 (53%) | 56 (54%) |
| Severe | 30 (26%) | 29 (28%) |
| Unknown | 2 (2%) | 1 (1%) |
| Seizures | 31 (27%) | 43 (41%) |
| Received therapeutic hypothermia | 60 (52%) | 50 (48%) |
| *Outcomes* |  |  |
| Death or major disability | 66 (57%) | 56 (61%), (n= 92) |
| Death | 35 (30%) | 34 (34%), (n= 101) |
| Major Disability | 31 (27%) | 22 (24%), (n= 92) |

In comparison to infants excluded in this analysis (Table 1), all baseline characteristics were similar except for a lower proportion of infants with seizures among included infants (27% vs 41%). The proportion of infants with death or major disability was also similar (57% vs 61%).

In the unadjusted analysis, infants near the normoxic range of pO_2_ (40-99 mmHg) had the lowest probability of death or disability in comparison to hypoxaemic and hyperoxaemic infants (Figure 1). The probability of death or disability plateaued at pO_2_ >200 mmHg. Excluding extreme outliers (pO_2_ >500 mmHg, n=5), the risk of death or disability in hyperoxaemic infants was 40/58 (0.69), which was higher than the risk in normoxaemic infants 20/48 (0.42).

**Figure 1.**
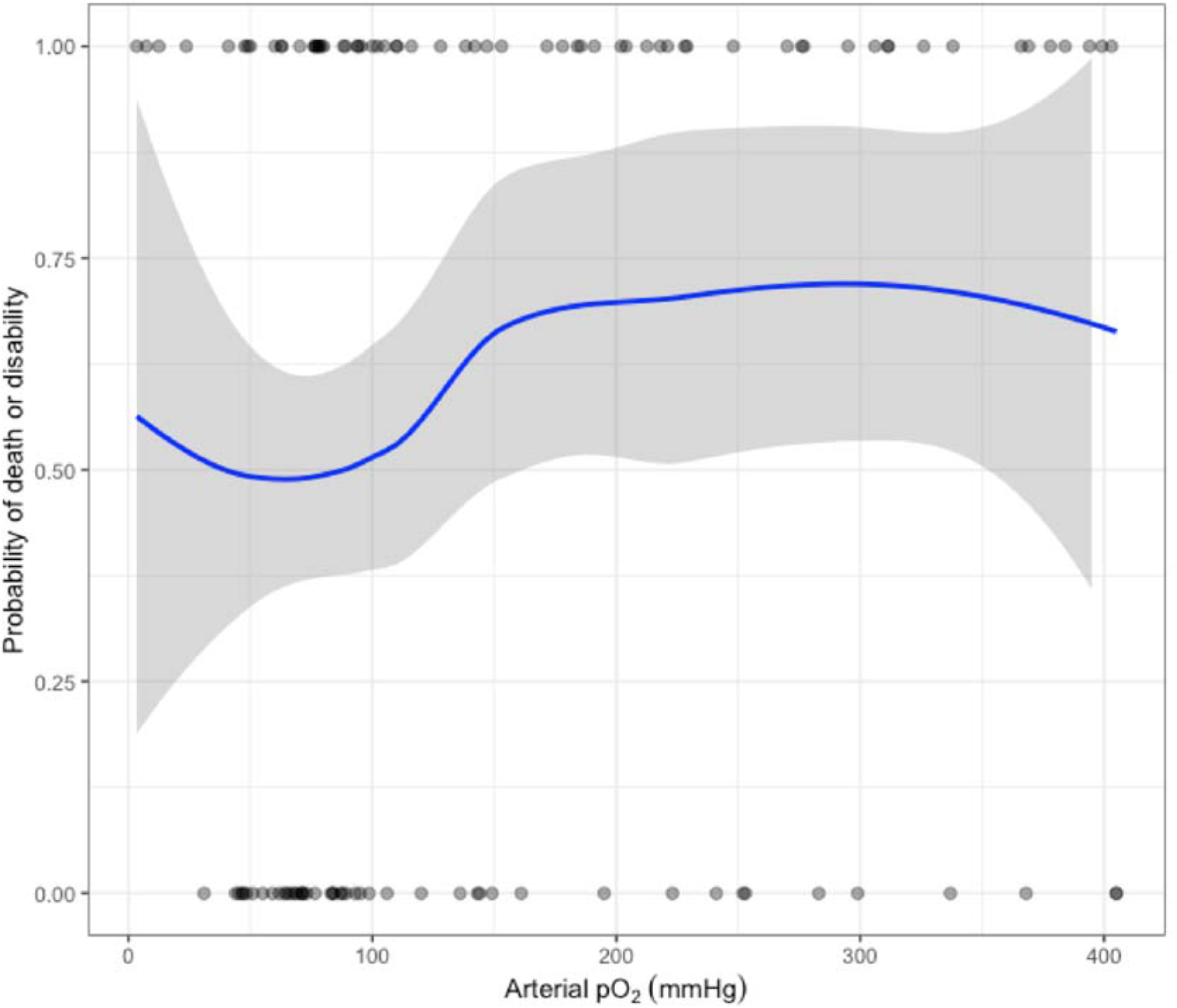
Loess curve showing the unadjusted probability (+/- 95% confidence interval) of death or major sensorineural disability in relation to arterial pO_2_ measured within 2 hours of birth. The yellow shaded area highlights the normoxaemic range (pO_2_ 40 – 99mmHg). Dots represent individual infants (n=111) who either did (plotted at y=1) or did not (plotted at y=0) have the study outcome. Extreme outliers with arterial pO_2_ >500mmHg were excluded (n=5).

A DAG model (Figure 2) informed the following minimally sufficient adjustment set of variables for estimating the causal effect of early hyperoxaemia with death or disability: pH, Apgar score at 10 min, time to first breath, chest compressions duration, adrenaline during cardiopulmonary resuscitation, first arterial pCO_2_. The univariable relationship with death or disability for each of these variables is shown in table 2.

**Table 2.** Univariable analysis of the relationship between the exposure of interest (hyperoxaemia) and the covariates identified for adjustment from the directed acyclic graph with the study outcome (death/disability). Infants with arterial pO_2_ <40mmHg (n= 5) and extreme outliers for pO_2_ (>500 mmHg, n=5) were excluded. ^a^ Data for pH was missing for 2 infants. ^b^ Data for Apgar score at 10 min was missing for 1 infant. Unadjusted risk ratios are for log transformed values. ^c^ Data for time to first breath was missing for 7 infants. Unadjusted risk ratios are for log transformed values. ^d^ Data for arterial pCO_2_ was missing for 2 infants. Unadjusted risk ratios are for log transformed values. ^e^ The adjusted estimate and p-value for hyperoxaemia was derived from a log-binomial model that included as covariates pH, Apgar at 10 min (log transformed), Time to first breath (log transformed), Adrenaline, and Arterial pCO_2_ (log transformed). Adjusted estimates for the covariates are not shown as they are not interpretable in a causal inference framework.

|  | **Infants who died or had major disability (n= 60)** | **Infants who survived without major disability (n= 46)** | **Unadjusted risk ratio (95% CI)** | **Adjusted risk ratio (95% CI)<sup>e</sup>** | **p-value from adjusted model<sup>e</sup>** |
| --- | --- | --- | --- | --- | --- |
| Hyperoxaemia (pO<sub>2</sub> ≥ 100mmHg), mmHg | 40 (67%) | 18 (39%) | 1.66 (1.21 – 2.00) | 1.61 (1.07 – 2.00) | 0.03 |
| pH, mean (SD)<sup>a</sup> | 6.88 (0.22) | 6.98 (0.20) | 0.23 (0.03 – 0.87) |  |  |
| Apgar at 10 min, median [IQR]<sup>b</sup> | 4 [2 – 5] | 4 [3 – 5] | 0.53 (0.24 – 0.90) |  |  |
| Time to first breath, min, median [IQR]<sup>c</sup> | 15 [8 – 25] | 7 [4 – 10] | 1.31 (1.15 – 1.45) |  |  |
| Adrenaline | 36 (60%) | 13 (28%) | 1.74 (1.31 – 2.06) |  |  |
| Arterial pCO<sub>2</sub>, mmHg, median [IQR]<sup>d</sup> | 31 [23 – 44] | 35 [28 – 42] | 0.74 (0.40 – 1.11) |  |  |

**Figure 2.**
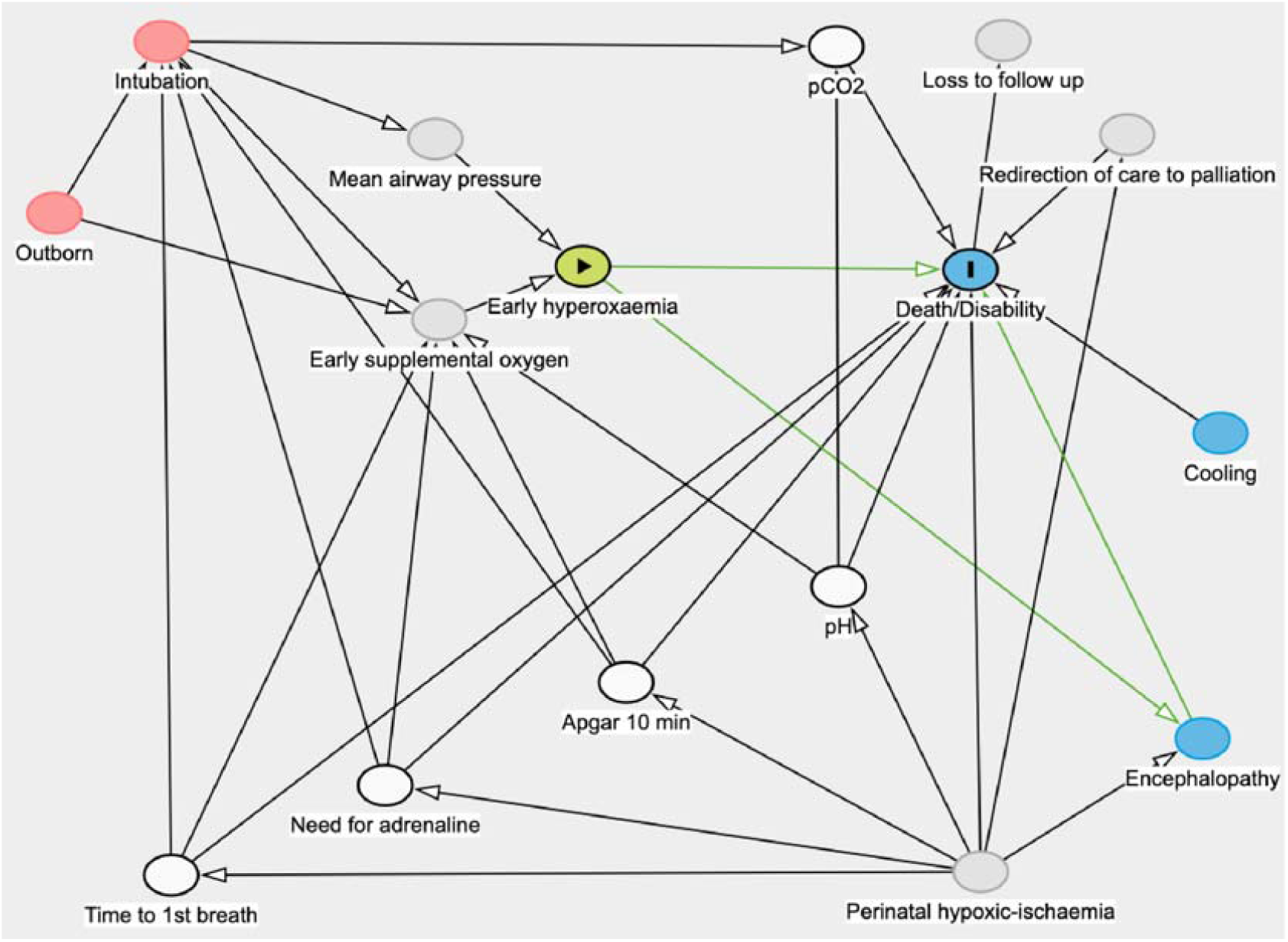
(A) Directed acyclic graph (DAG), with arrows used to indicate pathways between exposure (Early hyperoxaemia) and outcome (Death/disability). The green lines highlight the direct and indirect paths representing the causal relationship under investigation. The diagram includes covariates that are unmeasured (grey), ancestors of outcome (blue), and ancestors of exposure and outcome (red). A minimal sufficient adjustment set consisting of measured variables (white) were identified to examine the causal relationship between exposure and outcome.

In the regression model that included the covariates identified for adjustment from the directed acyclic graph, hyperoxaemia (admission arterial pO_2_ ≥ 100mmHg) was causally associated with an increased risk of death/disability (adjusted risk ratio 1.61, 95% CI 1.07 – 2.00, p= 0.03) in relation to normoxaemia (pO_2_ 40 – 99mmHg).

## Discussion

In this secondary causal analysis of data from infants in the ICE trial, we found that arterial hyperoxaemia within 2 hours of birth increased the risk of death or disability, following adjustment for severity of perinatal hypoxic-ischemia and pCO_2_. This finding is contingent upon sufficient adjustment for confounding, and assuming there are no residual causal or parametric biases.

Our findings are in keeping with Kapadia et al., who found in a cohort of 120 infants that the risk of developing moderate to severe HIE within 6 hours of birth was associated with arterial pO_2_ >100 mmHg within the first hour after birth.^14^ An early retrospective study from the pre-hypothermia era found an association between death or disability at 2 years of age and arterial pO_2_ >200 mmHg within 2 h of birth, severe hypocarbia and time to regular breathing.^15^ However, Pappas et al. in their analysis of data from the National Institute of Child Health and Human Development Neonatal Research Network trial of whole-body hypothermia found no univariable association between early hyperoxaemia and death/disability at 18 to 22 months of age.^16^ An important methodological limitation across these studies is the grouping together of hypoxaemic and normoxaemic infants into the comparator group. Given the U-shaped relationship that has also been described in the pediatric and adult literature,^2,19^ this approach would likely overestimate risk in the non-hyperoxaemic group and therefore underestimate the effect of hyperoxaemia. Comparing hyperoxaemia to normoxaemia addresses the clinical question of interest and increases the ability to detect an association if one exists.

Several pre-clinical studies lend biological plausibility to our findings. Dalen et al. found that in moderately asphyxic rats, the neuroprotective effect of therapeutic hypothermia was nearly fully negated by the increase in injury when using 100% oxygen for 30 minutes during reoxygenation when compared to room air.^20^ Koch et al. found that reoxygenation in 100% oxygen for 30 minutes caused accumulation of the oxidative metabolite nitrotyrosine, depleted preoligodendrocyte glial progenitors and impaired functional recovery of asphyxic mice when compared to reoxygenation in room air.^21^ Studies in piglets, however, found no differences in the neural injury marker S100 calcium binding protein B or other innate immune cytokines following 30 minute of exposure to 100% oxygen.^22,23^ More recently, we and others found in asphyxic lambs that marked exposure to cerebral hyperoxia occurs as early as in the first few minutes following return of circulation with current strategies of supplemental oxygen use.^9,24–26^

A strength in our study was the careful consideration of variables that may confound the relationship between early hyperoxaemia and death/disability. Clinicians providing advanced resuscitation to infants with severe perinatal hypoxic-ischemia are likely to use high concentrations of supplemental oxygen both during resuscitation and in the early stabilization phase. Exposure to hyperoxaemia may, however, directly contribute to cellular injury that leads to encephalopathy and death/disability. To better establish these relationships, we used a DAG to identify both confounders and mediators of the causal pathways between early hyperoxaemia and death/disability. The adjusted model therefore represents a best estimate of the causal relationship between early hyperoxaemia and death/disability in this dataset.

This study has important limitations. As with any non-randomized study, residual confounding from unmeasured or incompletely adjusted variables is likely to remain. The sample size is limited, particularly as the analysis was restricted to infants who had an arterial blood gas taken within 2 hours of birth. We were unable to estimate the relationship between hyperoxaemia and death/disability in infants who had a capillary or venous blood sample as the relationship with arterial pO_2_ is unknown. We were also unable to evaluate whether restricting inclusion to infants with an arterial sample resulted in important selection bias that affected the causal relationship between pO_2_ and death/disability. However, the baseline characteristics of excluded infants was very similar to infants included in the analysis (Table 1). The ICE trial was conducted during a period when the dangers of hyperoxia were less well recognized and oxygen is likely to have been more liberally used than in current practice. However, FiO_2_ >80% was an exclusion criterion for trial enrolment. Finally, we did not have data to calculate cumulative oxygen exposure. Partial pressure of oxygen over the first few hours after birth may better characterize oxygen exposure during the period when the brain is at highest risk of reperfusion injury. We are not aware of such oxygen exposure data collected in large studies of infants with HIE. Instead, for this analysis we relied on a cross-sectional measurement of arterial pO_2_ at the time of first blood gas analysis that may not be representative of overall oxygen burden. However, the requirement for an early blood gas (ideally within 1 h of birth) in the ICE study protocol meant that there was a degree of consistency in the timing of blood gas sampling.

At present, early hyperoxia is not considered in clinical practice to be an outcome modifier for death/disability in infants with HIE. Minimum core datasets, including HIE registries, may not currently include admission pO_2_ and specify route of collection.^27^ In contrast, the use of room air for commencing ventilation in infants with lesser degrees of perinatal hypoxic-ischemia (those not needing chest compressions) is now well established.^8^ The recognition that hyperoxia contributes to death and oxidative stress in these infants was confirmed in several quasi-randomized trials.^3^ However, no prospective studies evaluating the impact of hyperoxia have been conducted in infants with more severe degrees of perinatal hypoxic-ischemia. These infants are more susceptible to oxidative injury from depleted antioxidants, mitochondrial dysfunction and altered cerebral autoregulation.^28^ The findings from this exploratory study should encourage further work to confirm the relationship between hyperoxia and death/disability in the current era of routine therapeutic hypothermia and improved titration of supplemental oxygen. If excess oxygen exposure is indeed found to contribute to injury in infants with HIE, avoidance of hyperoxia could represent a simple and readily available way to improve outcomes for infants with HIE worldwide.

Strategies to avoid hyperoxia may require a reappraisal of how we target oxygenation in the immediate post-resuscitation phase. Following return of circulation, resuscitation recommendations provide either no clear guidance or recommend the targeting of preductal SpO_2_ levels observed in healthy infants.^8,29^ Recent animal studies have found that the immediate period following return of circulation is a critical window for limiting exposure to hyperoxia.^9,24–26^ This period is characterized by restoration of cardiac output to the lungs and a catecholamine-driven post-asphyxial surge of oxygenated blood to the brain. Historically, the focus has been on oxygen use *during* CPR, but blood flow to end-organs is minimal and hyperoxic exposure unlikely, irrespective of concentration of oxygen used.^9,25,26^ Beyond the delivery room, impaired cerebral autoregulation is well recognized in infants with HIE.^30^ The suppression of neuronal activity that characterizes the latent phase results in a lower threshold for excess cerebral oxygen delivery in relation to consumption. These lines of evidence provide a physiological basis for cerebral tissue hyperoxia despite oxygen saturation and pO_2_ levels that would be considered normal in well infants. Prospective studies evaluating lower or dynamic SpO_2_ targets, and/or incorporating the use of Near Infrared Spectroscopy, may be warranted if our findings in this study are replicated in larger and more recent datasets.

## Data Availability

All data produced in the present study are available upon reasonable request to the authors

## Acknowledgements

This study was supported by the National Health and Medical Research Council (NHMRC) through a Program Grant (#606789) and Fellowships (SBH: APP545921, GRP: APP1105526, PGD: APP1059111). SB was supported by an Australian Government Research Training Program Scholarship. The funders had no role in the in the study design, in the collection, analysis and interpretation of data; in the writing of the manuscript; and in the decision to submit the manuscript for publication.

## List of abbreviations

(SpO_2_): Arterial oxygen saturation
(BSID-II): Bayley Scales of Infant Development II
(CP): cerebral palsy
(CPR): cardiopulmonary resuscitation
(DAG): Directed acyclic graph
(FiO_2_): fraction of inspired oxygen
(HIE): hypoxic ischemic encephalopathy
(pO_2_): partial pressure of oxygen
(pCO_2_): partial pressure of carbon dioxide
(GMFCS): Gross Motor Function Classification System.

